# Association Between SGLT2 Inhibitor Use and Post-Contrast Acute Kidney Injury in Patients Undergoing Coronary Angiography: A Multicenter Cohort Study

**DOI:** 10.64898/2026.02.20.26346726

**Authors:** Alper Azak, MünevverGül Avşar, Gülay Koçak, Alper Koyuncuoğlu, Kadriye Kılıçkesmez, Özge Kama Başcı, Eyup Avcı

## Abstract

**Introduction:** Patients with type 2 diabetes mellitus (T2DM) are at increased risk of coronary artery disease and frequently undergo coronary angiography or percutaneous coronary intervention. Although risk factors for post-contrast acute kidney injury (PC-AKI) are well defined, effective preventive strategies remain limited.

**Methods:** This multicenter observational cohort study included 975 patients aged 18–75 years who underwent coronary angiography and/or percutaneous coronary intervention with iodinated contrast between June 2023 and June 2024. All patients received standardized intravenous hydration. Participants were grouped according to chronic sodium–glucose co-transporter-2 (SGLT2) inhibitor use (≥3 months). PC-AKI was defined as a ≥25% or ≥0.5 mg/dL increase in serum creatinine within 48–72 hours after contrast exposure.

**Results:** The mean age was 59.2 ± 11.7 years, and 70.8% were male; 16.9% were using SGLT2 inhibitors. PC-AKI occurred in 7.3% of patients, and 0.7% required renal replacement therapy. In univariate analysis, advanced age, diabetes, hypertension, heart failure, diuretic use, and elevated urea, creatinine, potassium, and uric acid levels were associated with PC-AKI. Higher eGFR, albumin, sodium levels, and SGLT2 inhibitor use were inversely associated. In multivariate analysis, age ≥65.5 years (OR 4.53), diabetes (OR 2.49), and uric acid >6.75 mg/dL (OR 2.34) remained independent risk factors, while eGFR >81.5 mL/min/1.73 m^2^ (OR 0.38), sodium >137.5 mmol/L (OR 0.36), and SGLT2 inhibitor use (OR 0.09) were independently protective.

**Conclusion:** Beyond established cardioprotective and renoprotective effects, SGLT2 inhibitors may reduce the risk of PC-AKI in patients with T2DM, potentially through decreased renal oxygen consumption and attenuation of contrast-induced hypoxic injury.

## 1. Introduction

Iodinated contrast media, widely utilized in a broad range of diagnostic imaging modalities and interventional procedures, have been extensively associated with the development of contrast-related renal dysfunction. ^1^ This adverse effect is particularly pronounced in individuals with pre-existing vulnerabilities, in whom contrast exposure may precipitate renal hemodynamic alterations, direct tubular toxicity, and inflammatory or oxidative pathways that collectively contribute to impaired kidney function. ^2^ Post-contrast acute kidney injury (PC-AKI) is generally defined as an impairment of renal function manifested by a ≥25% rise in serum creatinine from baseline or an absolute increase of 0.5 mg/dL (44 µmol/L) within 48– 72 hours after intravenous contrast administration. ^3^

PC-AKI remains a growing concern given the rising number of contrast-based coronary procedures performed in high-risk patients, such as those with chronic kidney disease (CKD), T2DM, or hypertension. ^4 5^ In these populations, a sudden decline in kidney function after angiography or intervention is a common complication, often attributable to PC-AKI. Although various risk factors for PC-AKI have been identified, effective preventive strategies (beyond adequate hydration) are limited. ^6^

Following administration, iodinated contrast media are rapidly distributed within the extracellular compartment and are cleared almost exclusively via glomerular filtration. ^7^ In patients with reduced baseline GFR, CM remains longer in circulation, and its intratubular concentration can increase up to 100-fold as water is reabsorbed in the tubules. ^8^ The pathogenesis of PC-AKI is multifactorial, involving renal ischemia (due to altered microcirculation) and direct tubular toxicity from the contrast agent. Animal studies have shown that contrast exposure can cause epithelial cell necrosis in the renal medulla (particularly in the thin ascending limb of Henle). The renal medulla is especially susceptible to hypoxic injury, and CM can exacerbate medullary hypoxia by diverting blood flow toward the renal cortex. Moreover, CM perturbs the balance of vasoactive mediators: it increases levels of renal vasoconstrictors (e.g., angiotensin II, endothelin, adenosine, vasopressin) while reducing vasodilator effects (e.g., nitric oxide, prostaglandins). This leads to renal vasoconstriction, hemoconcentration, and diminished medullary blood flow, thereby worsening tissue hypoxia and injury in kidney tubules. Clinically, PC-AKI is associated with prolonged hospitalization, higher readmission rates, and increased short- and long-term morbidity and mortality. CKD and T2DM are well-established risk factors for PC-AKI, and in patients with pre-existing CKD the incidence of PC-AKI has been reported to range from 10% to 20%. ^9^

Sodium–glucose co-transporter 2 (SGLT2), predominantly expressed in the proximal convoluted tubule, serves as the principal mediator of renal glucose reabsorption, accounting for approximately 80–90% of filtered glucose uptake. SGLT2 is primarily located in the proximal convoluted tubule, where it plays a central role in renal glucose handling by reclaiming approximately 80–90% of the filtered glucose load. ^10^ Beyond glycemic control, SGLT2 inhibitors confer additional benefits such as reductions in blood pressure, body weight, serum uric acid, liver steatosis, oxidative stress, and inflammation. Notably, large cardiovascular outcome trials have demonstrated significant reductions in cardiovascular events and renal endpoints with SGLT2 inhibitor therapy in patients with T2DM. Consequently, these agents have gained considerable attention as a promising therapeutic option, offering effective glycemic control while simultaneously conferring protective benefits to multiple organ systems. ^11 12 13 14^ The nephroprotective effects of SGLT2 inhibitors are believed to arise from a combination of hemodynamic and cellular effects, including reductions in intraglomerular pressure, improvements in renal oxygen balance, and attenuation of inflammatory and fibrotic processes within the kidney. ^15^

In this study, we aimed to investigate the association between SGLT2 inhibitor use and the development of PC-AKI in the overall population of patients undergoing coronary angiography and/or percutaneous coronary intervention. Additionally, we sought to explore whether the potential protective effect of SGLT2 inhibitors differed across clinically relevant subgroups, including patients at higher risk for PC-AKI.

## 2. Methods

### 2.1 Study Population and Design

This study was designed as a multicenter, observational cohort study conducted across three tertiary care centers from June 2023 through June 2024. Patients who underwent coronary angiography and/or percutaneous coronary intervention with iodinated contrast media during the study period were evaluated. Baseline demographic characteristics, comorbid conditions, medication use, and laboratory parameters were recorded prior to contrast exposure. Baseline renal function of eligible patients ranged from eGFR 25 to 115 mL/min/1.73 m^2^. All patients received standardized periprocedural hydration consisting of intravenous 0.9% saline administered at a rate of 1 mL/kg/hour for 6 hours prior to and 12 hours following the procedure, in accordance with institutional protocols for the prevention of contrast-related renal injury.

Patients were excluded if they had acute kidney injury at baseline or if they had received any prophylactic pharmacologic interventions for contrast nephropathy (such as N-acetylcysteine or sodium bicarbonate) prior to the procedure. We also excluded patients with incomplete medical records, those who received >100 mL of contrast volume or a contrast agent other than an iso-osmolar CM, and patients who could not tolerate aggressive hydration due to risk of volume overload (e.g. severe heart failure or pulmonary edema). The study protocol was approved by the ethics committee of University of Health Sciences Balikesir Ataturk Education and Research Hospital.

### 2.2 SGLT2 Inhibitor Exposure

Patients were classified as SGLT2 inhibitor users if they had been receiving an SGLT2 inhibitor for a minimum duration of three months prior to contrast exposure, a period considered sufficient to confer potential renoprotective effects. Individuals with type 2 diabetes mellitus who had no history of SGLT2 inhibitor use, or who had discontinued therapy more than three months before the procedure, were categorized as non-users. Based on SGLT2 inhibitor exposure status at the time of hospital admission, patients were subsequently assigned to either the user or non-user group

### 2.3 Primary Outcome

The primary outcome of interest was the development of PC-AKI. PC-AKI was defined as an impairment of renal function characterized by either a ≥25% increase in serum creatinine from baseline or an absolute increase of 0.5 mg/dL (44 µmol/L) in serum creatinine within 48–72 hours after exposure to contrast media, in the absence of alternative causes for acute kidney injury. This definition is consistent with standard criteria for contrast-induced nephropathy. Serum creatinine was measured at baseline (before contrast administration) and monitored for up to 3 days post-procedure to detect any changes meeting the above criteria.

### 2.4 Statistical Analysis

All statistical analyses were performed using SPSS version 24.0 (IBM Corp., Armonk, NY, USA) and Microsoft® Excel® 2019 (32-bit). Continuous variables are presented as mean ± standard deviation, and categorical variables are presented as frequencies or percentages. Receiver operating characteristic (ROC) curve analysis was performed to evaluate the predictive ability of continuous variables for the development of PC-AKI and to determine optimal cut-off values based on sensitivity and specificity. The area under the curve (AUC) with corresponding 95% confidence intervals was reported.

Univariate logistic regression analysis was used to identify variables associated with the development of PC-AKI. Variables demonstrating statistical significance in univariate analysis were subsequently entered into a multivariate logistic regression model using a forward stepwise selection method based on the likelihood ratio test to determine independent predictors of PC-AKI. Odds ratios (ORs) with 95% confidence intervals (CIs) were calculated. The final step of the stepwise procedure was reported as the definitive multivariate model. All statistical tests were two-sided, and a p value <0.05 was considered statistically significant. The Type I error rate was set at 5%. A formal a priori sample size calculation was not performed. However, a post hoc power analysis was conducted based on the observed effect size. With a total study population of 975 patients and 71 post-contrast acute kidney injury events, the study demonstrated adequate statistical power (>80%) to detect the observed association between SGLT2 inhibitor use and the risk of PC-AKI (adjusted OR ≈ 0.09) at a two-sided alpha level of 0.05.

## 3. Results

A total of 975 patients were included in the study. The median age of the patients was 59 (min:26–max:90) and the mean age was 59.2±11.7. Of the study population, 27.6% (n=269) were over 65 years of age and 70.8% (n=690) of the patients were male. Among the patients, 63.0% (n=614) had hypertension, 39% (n=380) had diabetes mellitus, and 15% (n=146) had congestive heart failure as a comorbidity. When the use of medications related to the renin– angiotensin system was evaluated, Angiotensin-converting enzyme (ACE) inhibitor use was detected at a rate of 28.8% (n=281) and angiotensin receptor blocker (ARB) use at 20.2% (n=197). The percentage of patients using SGLT2 inhibitors was 16.9% (n=165). The number of patients with an estimated glomerular filtration rate (eGFR) below 60 mL/min/1.73 m^2^ was 124 (7.3%), the number of patients who developed PC-AKI was 71 (7.3%), and renal replacement therapy requirement related to this was present in 7 patients (0.7%). Pre-contrast laboratory parameters were shown in Table 1 with mean and standard deviation values.

**Table 1.** Pre-Contrast Demographic and Laboratory Assessment of the Study Population (n = 975)

|  | N(%) |
| --- | --- |
| Age >65 years | 269 (27.6%) |
| Male sex | 690 (70.8%) |
| Diabetes mellitus | 380 (39.0%) |
| Hypertension | 614 (63.0%) |
| Congestive heart failure | 146 (15.0%) |
| SGLT2 inhibitor use | 165 (16.9%) |
| ARB use | 197 (20.2%) |
| ACE inhibitor use | 281 (28.8%) |
| Diuretic use | 333 (34.2%) |
| eGFR <60 mL/min/1.73 m<sup>2</sup> | 124 (12.7%) |
| PC-AKI development | 71 (7.3%) |
| Need for RRT | 7 (0.7%) |
| **Laboratory parameters** | **Mean value ± SD** |
| Urea (mg/dL) | 37.79 ± 17.23 |
| Creatinine (mg/dL) | 0.90 ± 0.29 |
| eGFR (mL/min/1.73 m<sup>2</sup>) | 87.81 ± 22.20 |
| Total protein (g/dL) | 6.83 ± 0.79 |
| Albumin (g/dL) | 4.06 ± 0.46 |
| Uric acid (mg/dL) | 5.50 ± 1.63 |
| Sodium (mmol/L) | 138.2 ± 4.53 |
| Potassium (mmol/L) | 4.47 ± 1.33 |
| Total cholesterol (mg/dL) | 175.72 ± 46.71 |
| LDL cholesterol (mg/dL) | 107.66 ± 40.04 |
| HDL cholesterol (mg/dL) | 41.49 ± 11.82 |
| Triglycerides (mg/dL) | 154.37 ± 118.84 |

**Abbreviations:** SD: Standard deviation, **SGLT2i:** Sodium–glucose cotransporter-2 inhibitor, **ARB:** Angiotensin receptor blocker, **ACE:** Angiotensin-converting enzyme, **eGFR:** Estimated glomerular filtration rate. **RRT:** Renal replacement therapy

The ideal cut-off values of the variables to predict post-contrast nephropathy were evaluated using ROC curve analysis. The ideal cut-off value for age was 65.5 years (AUC: 0.779; sensitivity: 69.6%; specificity: 74.7%; p<0.001), the cut-off value for urea was 46.5 mg/dL (AUC: 0.756; sensitivity: 56.5%; specificity: 84.5%; p<0.001), the cut-off value for serum creatinine was determined as 0.94 mg/dL (AUC: 0.757; sensitivity: 71.7%; specificity: 66.7%; p<0.001), and the cut-off value for eGFR was calculated as 81.5 mL/min/1.73 m^2^ (AUC: 0.774; sensitivity: 65.3%; specificity: 83.7%; p<0.001). The cut-off value for serum albumin was 3.92 g/dL (AUC: 0.719; sensitivity: 70.1%; specificity: 67.3%; p=0.001), for uric acid was 6.75 mg/dL (AUC: 0.674; sensitivity: 56.5%; specificity: 78.8%; p=0.006), and for sodium was determined as 137.5 mmol/L (AUC: 0.655; sensitivity: 63.5%; specificity: 61.2%; p<0.001). No ideal cut-off could be determined for total protein (p=0.152), potassium (p=0.062), total cholesterol (p=0.932), LDL cholesterol (p=0.924), HDL cholesterol (p=0.890), and triglyceride (p=0.865), and median values were used in regression analyses. (Table 2) (Figure 1).

**Table 2.** Receiver Operating Characteristic (ROC) Curve Analysis of Continuous Variables for the Prediction of Post-Contrast Acute Kidney Injury.

| Variable | AUC | AUC 95% CI | p-value | Cut-off | Sens. (%) | Spec. (%) |
| --- | --- | --- | --- | --- | --- | --- |
| Age (years) | 0.779 | 0.682–0.877 | **<0.001** | 65.5 | 69.6 | 74.7 |
| Urea (mg/dL) | 0.756 | 0.657–0.854 | **<0.001** | 46.5 | 56.5 | 84.5 |
| Creatinine (mg/dL) | 0.757 | 0.651–0.862 | **<0.001** | 0.94 | 71.7 | 66.7 |
| eGFR (mL/min/1.73 m<sup>2</sup>) | 0.774* | 0.711–0.836 | **<0.001** | 81.5 | 65.3 | 83.7 |
| Total protein (g/dL) | 0.409 | 0.283–0.535 | 0.152 | - | - | - |
| Albumin (g/dL) | 0.719* | 0.642–0.795 | **0.001** | 3.92 | 70.1 | 67.3 |
| Uric acid (mg/dL) | 0.674 | 0.552–0.795 | 0.006 | 6.75 | 56.5 | 78.8 |
| Sodium (mmol/L) | 0.655* | 0.570–0.741 | **<0.001** | 137.5 | 63.5 | 61.2 |
| Potassium (mmol/L) | 0.618 | 0.466–0.771 | 0.062 | - | - | - |
| Total cholesterol (mg/dL) | 0.495 | 0.367–0.622 | 0.932 | - | - | - |
| LDL cholesterol (mg/dL) | 0.494 | 0.363–0.625 | 0.924 | - | - | - |
| HDL cholesterol (mg/dL) | 0.509 | 0.396–0.621 | 0.890 | - | - | - |
| Triglycerides (mg/dL) | 0.511 | 0.376–0.646 | 0.865 | - | - | - |

**Abbreviations:** **Sens:** Sensitivity **Spec:** Specificity , **ROC:** Receiver operating characteristic, **AUC:** Area under the curve, **CI:** Confidence interval, **eGFR:** Estimated glomerular filtration rate.
\*ROC curves were adjusted by inverting the direction where app

**Figure 1.**
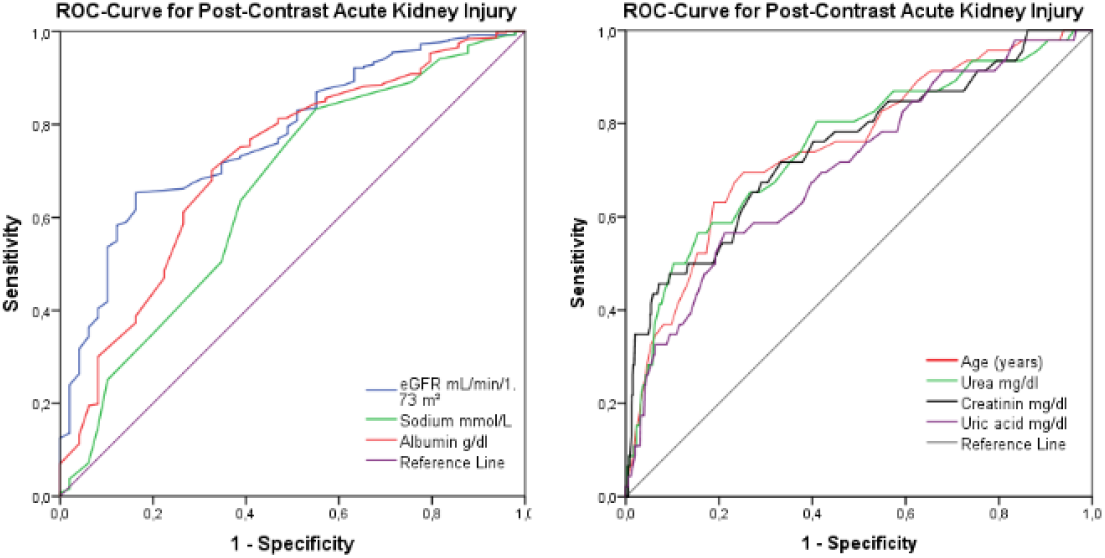
ROC-Curve showing acute kidney injury and curves for statistically significant variables.

Factors predicting post-contrast acute kidney injury were evaluated using univariate logistic regression analysis. Age ≥65.5 years (OR: 4.61; 95% CI: 2.80–7.59; p<0.001), presence of diabetes mellitus (OR: 2.01; 95% CI: 1.24–3.27; p=0.005), presence of hypertension (OR: 2.32; 95% CI: 1.29–4.16; p=0.005), congestive heart failure (OR: 1.89; 95% CI: 1.06–3.37; p=0.030), diuretic use (OR: 2.38; 95% CI: 1.46–3.87; p<0.001), urea >46.5 mg/dL (OR: 4.97; 95% CI: 3.02–8.17; p<0.001), creatinine >0.94 mg/dL (OR: 3.65; 95% CI: 2.22–6.02; p<0.001), potassium >4.4 mmol/L (OR: 1.84; 95% CI: 1.12–3.03; p=0.016), and uric acid >6.75 mg/dL (OR: 4.82; 95% CI: 2.62–8.89; p<0.001) were identified as factors increasing the risk of contrast nephropathy. eGFR >81.5 mL/min/1.73 m^2^ (OR: 0.15; 95% CI: 0.09–0.25; p<0.001), albumin >3.92 g/dL (OR: 0.22; 95% CI: 0.12–0.40; p<0.001), sodium >137.5 mmol/L (OR: 0.44; 95% CI: 0.27–0.71; p=0.001), and SGLT2 inhibitor use (OR: 0.28; 95% CI: 0.10–0.77; p=0.014) were evaluated as risk-reducing factors for contrast nephropathy (Table 3) (Figure 2).

**Table 3.**
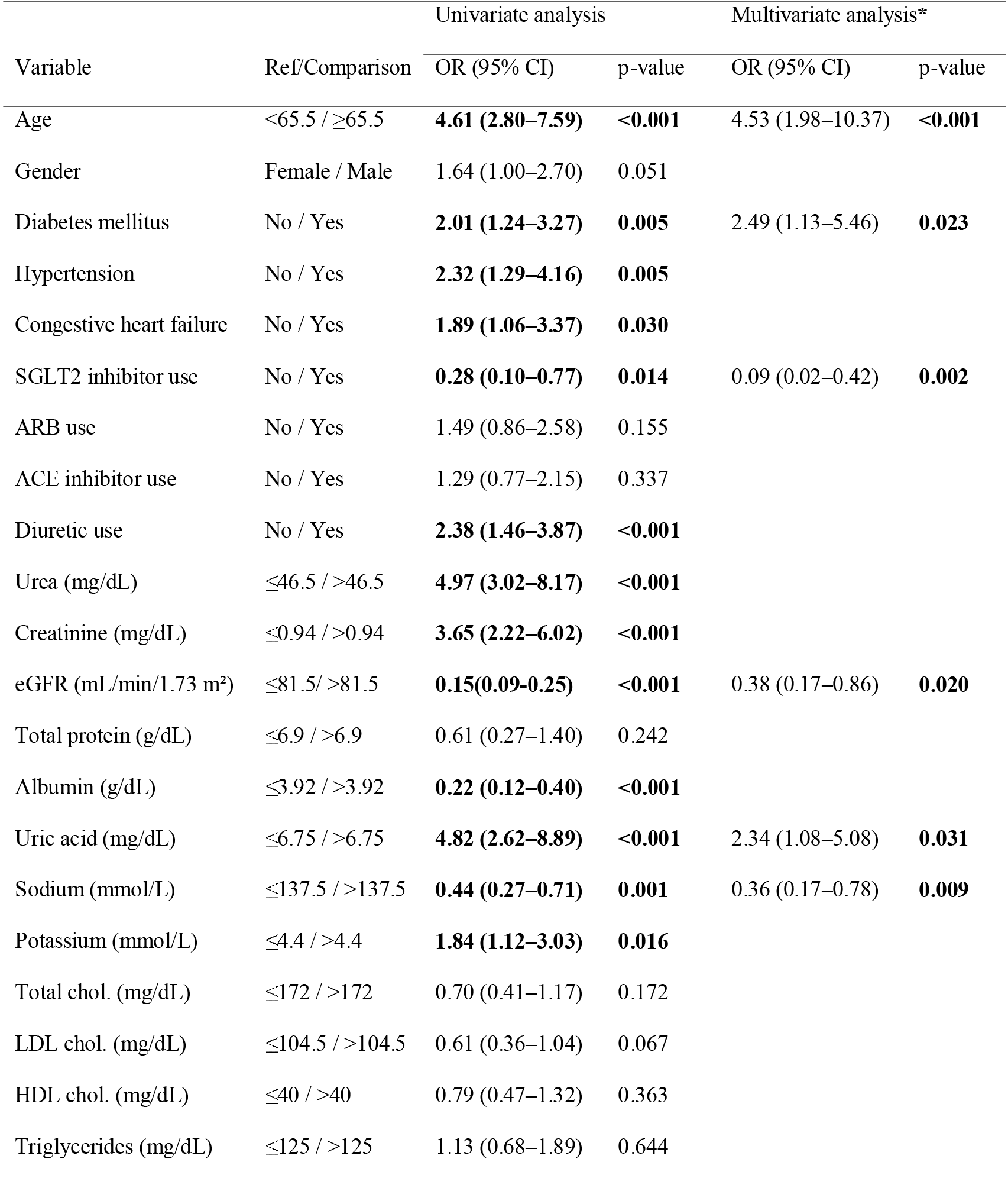
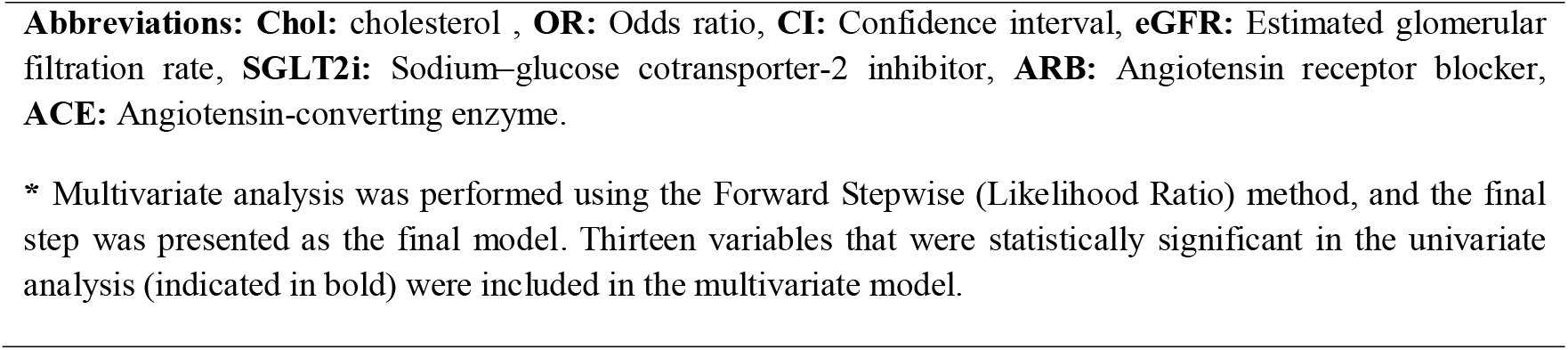
Univariate and Multivariate Logistic Regression Analyses of Factors Predicting Post-Contrast Acute Kidney Injury.

| Variable | Ref/Comparison | Univariate analysis |  | Multivariate analysis* |  |
| --- | --- | --- | --- | --- | --- |
|  |  | OR (95% CI) | p-value | OR (95% CI) | p-value |
| Age | <65.5 / ≥65.5 | **4.61 (2.80–7.59)** | **<0.001** | 4.53 (1.98–10.37) | **<0.001** |
| Gender | Female / Male | 1.64 (1.00–2.70) | 0.051 |  |  |
| Diabetes mellitus | No / Yes | **2.01 (1.24–3.27)** | **0.005** | 2.49 (1.13–5.46) | **0.023** |
| Hypertension | No / Yes | **2.32 (1.29–4.16)** | **0.005** |  |  |
| Congestive heart failure | No / Yes | **1.89 (1.06–3.37)** | **0.030** |  |  |
| SGLT2 inhibitor use | No / Yes | **0.28 (0.10–0.77)** | **0.014** | 0.09 (0.02–0.42) | **0.002** |
| ARB use | No / Yes | 1.49 (0.86–2.58) | 0.155 |  |  |
| ACE inhibitor use | No / Yes | 1.29 (0.77–2.15) | 0.337 |  |  |
| Diuretic use | No / Yes | **2.38 (1.46–3.87)** | **<0.001** |  |  |
| Urea (mg/dL) | ≤46.5 / >46.5 | **4.97 (3.02–8.17)** | **<0.001** |  |  |
| Creatinine (mg/dL) | ≤0.94 / >0.94 | **3.65 (2.22–6.02)** | **<0.001** |  |  |
| eGFR (mL/min/1.73 m<sup>2</sup>) | ≤81.5 / >81.5 | **0.15(0.09–0.25)** | **<0.001** | 0.38 (0.17–0.86) | **0.020** |
| Total protein (g/dL) | ≤6.9 / >6.9 | 0.61 (0.27–1.40) | 0.242 |  |  |
| Albumin (g/dL) | ≤3.92 / >3.92 | **0.22 (0.12–0.40)** | **<0.001** |  |  |
| Uric acid (mg/dL) | ≤6.75 / >6.75 | **4.82 (2.62–8.89)** | **<0.001** | 2.34 (1.08–5.08) | **0.031** |
| Sodium (mmol/L) | ≤137.5 / >137.5 | **0.44 (0.27–0.71)** | **0.001** | 0.36 (0.17–0.78) | **0.009** |
| Potassium (mmol/L) | ≤4.4 / >4.4 | **1.84 (1.12–3.03)** | **0.016** |  |  |
| Total chol. (mg/dL) | ≤172 / >172 | 0.70 (0.41–1.17) | 0.172 |  |  |
| LDL chol. (mg/dL) | ≤104.5 / >104.5 | 0.61 (0.36–1.04) | 0.067 |  |  |
| HDL chol. (mg/dL) | ≤40 / >40 | 0.79 (0.47–1.32) | 0.363 |  |  |
| Triglycerides (mg/dL) | ≤125 / >125 | 1.13 (0.68–1.89) | 0.644 |  |  |

**Abbreviations:** **Chol:** cholesterol , **OR:** Odds ratio, **CI:** Confidence interval, **eGFR:** Estimated glomerular filtration rate, **SGLT2i:** Sodium–glucose cotransporter-2 inhibitor, **ARB:** Angiotensin receptor blocker, **ACE:** Angiotensin-converting enzyme.
\* Multivariate analysis was performed using the Forward Stepwise (Likelihood Ratio) method, and the final step was presented as the final model. Thirteen variables that were statistically significant in the univariate analysis (indicated in bold) were included in the multivariate model.

**Figure 2.**
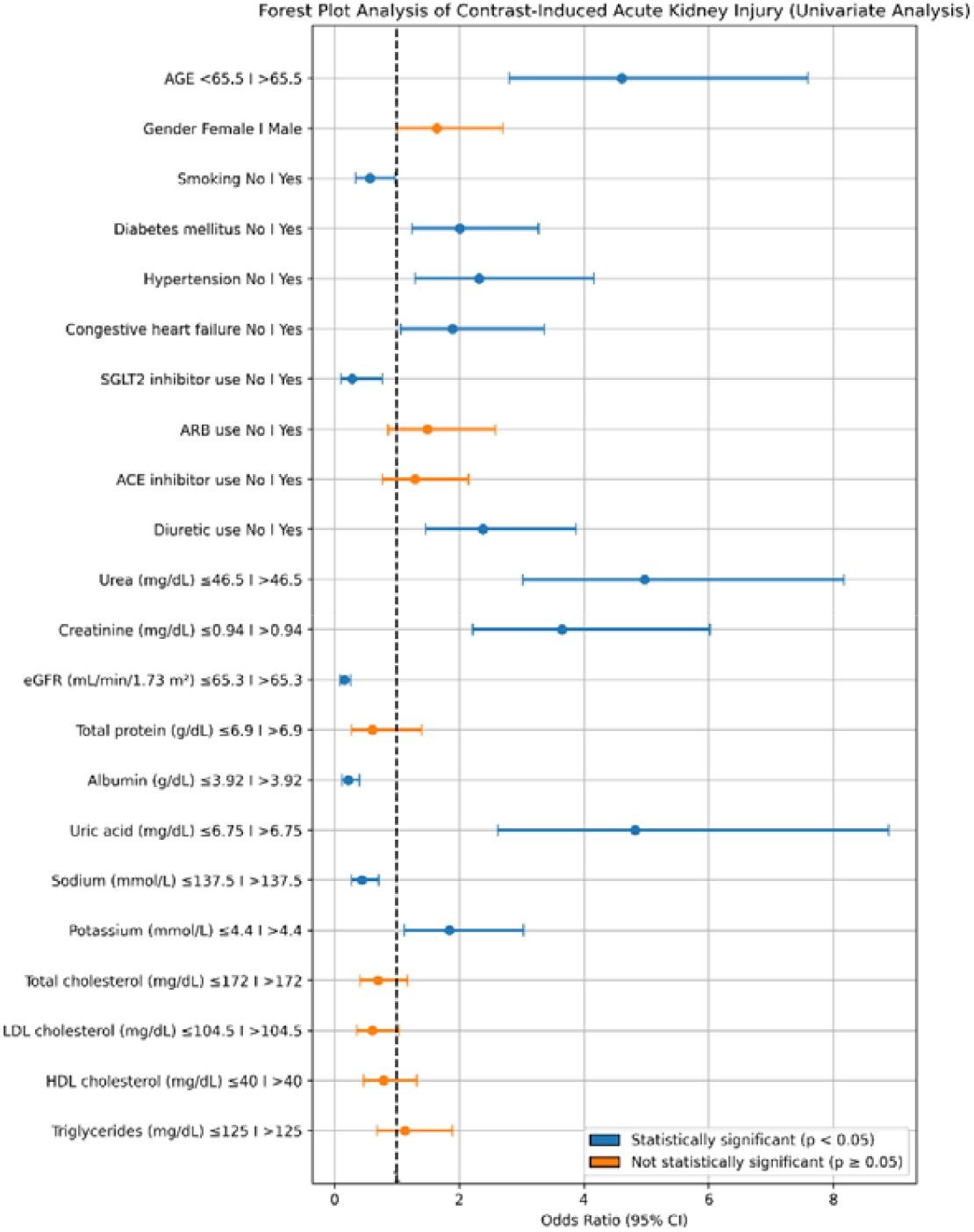
Forest plot of univariate logistic regression analysis of factors affecting acute kidney injury.

Thirteen variables that were statistically significant in univariate analysis were analyzed using multivariate logistic regression analysis with the Forward-LR stepwise method, and six variables constituted a predictive model. According to the predictive model, age ≥65.5 years constituted approximately a 4-fold risk (OR: 4.06; 95% CI: 1.77–9.31; p=0.001), presence of DM approximately a 2.5-fold risk (OR: 2.49; 95% CI: 1.13–5.46; p=0.023), and uric acid level >6.75 approximately a 2.3-fold risk (OR: 2.34; 95% CI: 1.08–5.08; p=0.031) for contrast nephropathy. eGFR >81.5 mL/min/1.73 m^2^ (OR: 0.38; 95% CI: 0.17–0.86; p=0.020), sodium level >137.5 (OR: 0.36; 95% CI: 0.17–0.78; p=0.009), and SGLT2 use (OR: 0.09; 95% CI: 0.02–0.42; p=0.002) were modeled as risk-reducing factors (Table 3).

## 4. Discussion

In this observational multicenter study, factors predicting post-contrast acute renal injury (PC-AKI) were investigated using data from 975 patients who experienced contrast exposure due to coronary angiography or percutaneous coronary intervention. In univariate logistic regression analysis; Patients aged ≥65.5 years (OR=4.61, p<0.001), presence of diabetes mellitus (OR=2.01, p=0.005), hypertension (OR=2.32, p=0.005), congestive heart failure (OR=1.89, p=0.030), diuretic use (OR=2.38, p<0.001), urea level >46.5 mg/dL (OR=4.97, p<0.001), creatinine level >0.94 mg/dL (OR=3.65, p<0.001), potassium level >4.4 mmol/L (OR=1.84, p=0.016), and uric acid level >6.75 mg/dL (OR=4.82, p<0.001) were identified as risk factors associated with the development of PC-AKI. In contrast, eGFR >81.5 mL/min/1.73 m^2^ (OR=0.15, p<0.001), albumin level >3.92 g/dL (OR=0.22, p<0.001), sodium level >137.5 mmol/L (OR=0.44, p=0.001), and SGLT2 inhibitor use (OR=0.28, p=0.014) were found to be associated with a decreased risk of PC-AKI. In multivariate analysis, being 65 years of age or older (OR=4.53, p<0.001), the presence of diabetes mellitus (OR=2.49, p=0.023), and uric acid level >6.75 mg/dL (OR=2.34, p=0.031) were identified as independent predictors associated with increased risk factors for the development of PC-AKI; An eGFR >81.5 mL/min/1.73 m^2^ (OR=0.38, p=0.020), sodium levels >137.5 mmol/L (OR=0.36, p=0.009), and SGLT2 inhibitor use (OR=0.09, p=0.002) were identified as independent predictive factors reducing the risk of PC-AKI.

Contrast-induced acute kidney injury (PC-AKI) remains a significant clinical challenge, particularly in patients with type 2 diabetes mellitus (T2DM), due to underlying microvascular dysfunction and associated comorbidities.^16^ Evaluating the effectiveness of protective approaches in this patient group is crucial for guiding clinical practice. The overall incidence of PC-AKI in our hydrated T2DM cohort (7.3%) was lower than historical rates reported in similar patients without prophylactic hydration. Previous studies have documented PC-AKI rates as high as 12–26% in high-risk T2DM patients who did not receive adequate periprocedural hydration. ^17 18 19^ This underscores the importance of volume expansion with IV fluids as a preventive measure for contrast nephropathy. Nonetheless, even with hydration, certain patients still developed PC-AKI, highlighting the need for additional protective strategies in vulnerable groups.

Contrast-induced renal injury occurs through a combination of hemodynamic and direct toxic effects, as discussed earlier. The administration of CM triggers alterations in renal perfusion and oxygen utilization that can injure susceptible kidney regions. Animal models have demonstrated that contrast can cause selective necrosis of tubular cells in the outer medulla (particularly the thick ascending limb), likely due to intense vasoconstriction and hypoxic injury in that region. ^20^ The medullary ischemia is partly caused by contrast-mediated shifts in blood flow from the medulla to the cortex. ^21^ Additionally, contrast media cause an imbalance in vasoactive substances: they raise intrarenal levels of vasoconstrictors (such as angiotensin II, endothelin, vasopressin, and adenosine) while reducing vasodilatory forces (like nitric oxide and prostaglandins). ^22 23^ This imbalance leads to reduced renal blood flow, especially in the medulla, and promotes hemoconcentration, further worsening hypoxia in renal tissues. ^24^

These pathophysiological insights explain why patients with pre-existing renal impairment or diabetes (who may already have endothelial dysfunction and microvascular disease) are at heightened risk for PC-AKI. Indeed, PC-AKI is not only a common iatrogenic complication but is also linked to worse clinical outcomes, including prolonged hospitalization and increased mortality. ^25^ Recognizing risk factors such as CKD and T2DM is therefore crucial in clinical practice, and guidelines recommend vigilant preventive measures in these high-risk groups. ^26^

SGLT2 inhibitors stand out as an important class of treatment that provides significant benefits to cardiovascular and renal health in diabetic patients.^27^ These agents inhibit glucose and sodium reabsorption in the proximal tubule, leading to glucosuria and natriuresis.^28^ The resulting osmotic diuresis and sodium excretion contribute to a decrease in blood pressure and reduced intravascular volume load. More importantly, inhibition of sodium reabsorption in the proximal tubule increases the amount of sodium reaching the distal nephron, strengthening the tubuloglomerular feedback mechanism.^29^ This effect causes vasoconstriction in afferent arterioles, reduces glomerular hyperfiltration, and normalizes intraglomerular pressure, contributing to the preservation of glomerular structure. ^2 30^ Through these mechanisms, SGLT2 inhibitors decrease renal oxygen consumption (the workload of sodium reabsorption is energetically costly) and improve renal cortical oxygenation. Recent studies also suggest that SGLT2 inhibition may directly ameliorate hypoxia-induced renal injury at the molecular level. In a murine model, dapagliflozin attenuated contrast-induced AKI by suppressing the hypoxia-inducible factor-1α (HIF-1α)/HE4/NF-κB signaling pathway, which is implicated in inflammatory and fibrotic responses to hypoxic stress. This finding provides a mechanistic link between SGLT2 inhibitor action and protection against contrast media toxicity in the kidney. ^31^

Another important consideration is the safety profile of SGLT2 inhibitors concerning AKI. Early after their introduction, there were case reports raising concern that SGLT2 inhibitors might precipitate AKI in some patients (possibly due to volume depletion or hypotension). However, most reported cases of SGLT2 inhibitor–associated AKI were reversible, and patients recovered renal function. ^32^ The consensus from larger studies and clinical trials is that SGLT2 inhibitors do not increase the risk of AKI; in fact, they may reduce it. Major outcome trials of dapagliflozin and empagliflozin in diabetes and CKD populations have shown cardio-renal benefits without any excess of AKI events compared to placebo. ^11 12 13 14^ Some analyses have even reported a lower incidence of AKI in patients on SGLT2 inhibitors versus other glucose-lowering therapies. ^33^ Our results align with these observations. We found that T2DM patients on chronic SGLT2 inhibitor therapy had significantly fewer episodes of PC-AKI than those not on SGLT2 inhibitors. This protective association remained evident even after accounting for differences in baseline characteristics, suggesting a potential renoprotective effect of SGLT2 inhibitors in the acute setting of contrast exposure.

Our findings are supported by other emerging clinical data. Özkan et al. reported that among diabetic patients with non-ST elevation myocardial infarction undergoing angiography, those receiving SGLT2 inhibitors had a lower rate of contrast-induced nephropathy than those not on SGLT2 inhibitors. ^34^ Similarly, a recent propensity-matched analysis by Liu *et al*. found that dapagliflozin use was associated with a significantly reduced risk of PC-AKI in T2DM patients with CKD undergoing elective coronary procedures. ^35^ These studies, together with our own, suggest that the nephroprotective properties of SGLT2 inhibitors can translate into fewer AKI events in the context of iodinated contrast exposure.

There are several possible explanations for why SGLT2 inhibitors confer protection in this setting. By inducing natriuresis and osmotic diuresis, SGLT2 inhibitors ensure better volume status and potentially prevent drastic drops in renal perfusion pressure during contrast administration. Their effect on reducing glomerular hyperfiltration could also mitigate the intraglomerular hypertension and high single-nephron GFR that predispose kidneys to injury. On a cellular level, lower renal tubular workload and improved oxygenation mean the kidney is more resilient to the hypoxic and oxidative stress caused by contrast media. Lastly, SGLT2 inhibitors have anti-inflammatory effects which might reduce the inflammatory component of contrast-induced nephropathy.

This study has some limitations. First, patients using SGLT2 inhibitors may differ in terms of unmeasurable variables such as general health status or the nature of clinical care they receive, and this may have affected the results. Although baseline variables such as age, baseline estimated glomerular filtration rate (eGFR), and concomitant comorbidities were controlled for with multivariate analyses, randomized controlled trials are needed to causally establish the protective effect. Furthermore, data on the specific types and doses of SGLT2 inhibitors used are not available, and long-term renal outcomes beyond the acute phase were not evaluated. Despite these limitations, the relatively large sample size and reliance on multicenter real-world data make our study strong. Additionally, the findings comparing the outcomes of post-contrast acute renal injury in patients using and not using SGLT2 inhibitors contribute to the limited literature in this area and encourage prospective studies by establishing a hypothesis that SGLT2 inhibitors may have a potential protective role against contrast-induced acute renal injury.

## 5. Conclusion

In T2DM, chronic hyperglycemia leads to upregulation of SGLT2 expression and increased reabsorption of sodium and glucose in the proximal tubule, which substantially increases renal oxygen consumption and contributes to medullary hypoxia. SGLT2 inhibitors, by blocking this pathway, reduce tubular workload and overall renal oxygen demand. In addition to their well-established long-term benefits in reducing cardiovascular events and slowing the progression of chronic kidney disease, SGLT2 inhibitors also appear to confer acute nephroprotection in settings of iodinated contrast exposure.

Our study demonstrates that prior use of SGLT2 inhibitors is associated with a significantly lower risk of PC-AKI in diabetic patients undergoing coronary interventions. This protective effect is likely multifactorial, arising from improvements in renal hemodynamics, enhanced renal oxygenation, and attenuation of inflammatory pathways. Taken together, these findings support consideration of SGLT2 inhibitors as part of a comprehensive renoprotective strategy for high-risk patients, although prospective randomized trials are still required to establish causality and define optimal peri-procedural management around contrast exposure.

## Data Availability

All data produced in the present work are contained in the manuscript. The data underlying this article will be shared upon reasonable request by the corresponding author.

https://zenodo.org/records/17229135/files/2duzeltilmis%20SGLT_FINALdata.sav?download=1

## Author Contributions

All authors were involved in conception, drafting, critical revision, and approval of the manuscript. The authors declare no conflicts of interest.

## Data availability statement

The data underlying this article will be shared upon reasonable request by the corresponding author.

## Ethics approval statement

Ethical approval for this research was granted by the University of Health Sciences Balikesir Ataturk Research and Education Hospital Ethics Committee during the meeting held on March 5, 2021 (approval number: 112).

## Patient consent statement

All patients are informed and written informed consent was obtained from all participants.

